# Three-dimensional cranial ultrasound and functional near infrared spectroscopy for bedside monitoring of intraventricular hemorrhage in preterm neonates

**DOI:** 10.1101/2022.10.16.22280949

**Authors:** Lilian M N Kebaya, Kevin Stubbs, Marcus Lo, Sarah Al-Saoud, Bradley Karat, Keith St Lawrence, Sandrine de Ribaupierre, Emma G. Duerden

## Abstract

Germinal Matrix-Intraventricular hemorrhage (GMH-IVH) remains a significant cause of adverse neurodevelopment in preterm infants. Current management relies on 2-dimensional cranial ultrasound (2D cUS) ventricular measurements. Reliable biomarkers are needed to aid in the early detection of posthemorrhagic ventricular dilatation (PHVD) and subsequent neurodevelopment. In a prospective cohort study, we incorporated 3-dimensional (3D) cUS and functional infrared spectroscopy (fNIRS) to monitor neonates with GMH-IVH. Preterm neonates (<32 weeks’ gestation) were enrolled following a GMH-IVH diagnosis. Neonates underwent sequential measurements: 3D cUS images were manually segmented using in-house software, and the ventricle volumes (VV) were extracted. Multichannel fNIRS data were acquired using a high-density system, and spontaneous functional connectivity (sFC) was calculated. Of the 30 neonates enrolled in the study, 21 (70%) had grade I-II and 12 (40%) grade III-IV GMH-IVH, and 23 neonates (77%) underwent surgical interventions to divert cerebrospinal fluid (CSF). Infants with severe GMH-IVH who underwent CSF diversion had larger VV and significantly decreased sFC (p<0.001). Our findings of increased VV and reduced sFC suggest that regional disruptions of ventricular size may impact the development of the underlying grey matter. Hence, 3D cUS and fNIRS are promising bedside tools for monitoring the progression of GMH-IVH in preterm neonates.

## Introduction

Germinal matrix-intraventricular hemorrhage (GMH-IVH) is the most common neurological complication faced by preterm born neonates (≤ 32 weeks gestation), affecting 25% of very low birth weight (VLBW) neonates (weighing < 1500g). (1-3) Preterm birth, defined as a live birth occurring ≤ 37 completed weeks of gestation (GA), remains the most critical risk factor for the development of GMH-IVH. (4, 5) Yearly, 15 million babies are born early, accounting for 11% of births worldwide and 8% in Canada. (6-8) Advancements in obstetrical and neonatal care have led to improved survival of preterm neonates; (9-16) especially the extremely low birth weight (ELBW) neonates, who are most affected by GMH-IVH (incidence of 45%). (17, 18) Hence, despite improved survival of preterm infants, there has been an increase in severe forms of GMH-IVH in ELBW neonates, which is associated with a high risk of death and neurodevelopmental impairment (NDI). (19, 20)

Preterm infants with GMH-IVH face complications: posthemorrhagic ventricular dilatation (PHVD) in the short term, cerebral palsy (CP), hearing and vision deficits, and cognitive and learning disabilities in the long term. (21-24) Preterm infants are susceptible to GMH-IVH due to immature germinal matrix (GM), maternal conditions, impaired autoregulation, alongside changes in blood flow. (25-29) Moreover, the first 72 h to 1 week after birth presents a delicate period of transition that ultimately predisposes these infants to complications, coinciding with the timing of GMH-IVH. (20, 30-32) Hence, routine cranial two-dimensional ultrasound (2D cUS) examination is recommended for all neonates ≤ 32 weeks GA within one week of life to identify and grade GMH-IVH; and subsequently at one month of age and term equivalent age (TEA) to identify white matter injury (WMI). (33, 34) cUS imaging is preferred due to its bedside availability, affordability and lack of ionizing radiation. cUS image quality, however, is dependent on the operator. Another cUS limitation is its inaccuracy in predicting long-term outcomes: mild GMH-IVH linked to NDI and severe IVH – good outcomes. Inconsistencies were attributed to the inability of cUS to take into account subtle WMI and in the latter - site and extent of the lesion. (35, 36) Lastly, cUS is limited in predicting sensorineural deficits and learning disabilities – equally important problems in the preterm neonate. Compared to cUS, MRI has the advantage of providing more detail to quantify and grade IVH and other complications. However, MRI use is limited by its high cost and the need for expertise in interpretation. Similarly, the need for sedation and transport of sick neonates makes MRI less ideal. (37, 38) Bedside tools are therefore needed to monitor hemorrhagic dilation of the lateral ventricles and changes in cerebral hemodynamics in the preterm neonate.

A promising bedside tool to monitor cerebral hemodynamics in preterms in the setting of the NICU is functional near-infrared spectroscopy (fNIRS). (38) This brain imaging technique is comparable to fMRI because it is sensitive to changes in blood oxygenation levels, an indirect marker of neuronal activity. Like fMRI, fNIRS can be used to calculate functional connectivity, a key measure of regional brain health. However, recent advances in fNIRS have overcome several limitations of neonatal MRI, in that fNIRS is portable, relatively insensitive to motion and has an excellent temporal resolution.

Within 1 to 3 weeks of GMH-IVH, 25% of GMH-IVH cases progress to PHVD - seen on serial cUS as increasing dilatation of ventricles. (17) Worsening PHVD leads to decreased blood flow to the cortex and WMI. (39) Of note, neonates with progressive PHVD, who require surgical interventions, may be at most risk for adverse brain and developmental outcomes. In fact, 35% of infants with progressive PHVD require surgical treatment. Additionally, mortality remains high in this population. (40) Long-term, PHVD is associated with NDI. (41) Management of progressive PHVD includes temporizing measures, such as lumbar taps, ventricular taps (VT), Ommaya reservoirs or external ventricular drains (EVD), and subsequently, a permanent shunt placement, if needed. However, there is little consensus regarding the timing of intervention for PHVD (42), with proponents for an earlier approach while others encourage a later timed surgical approach. (43) Despite extensive research, managing PHVD remains challenging in the preterm neonate.

Precise measurement of the degree of PHVD to guide timely intervention is vital. Ongoing research has shown the utility of three-dimensional cranial US (3D cUS) volumetric thresholds to guide intervention. (44) A follow-up study of infants with PHVD showed that ventricular volume (VV) affected outcomes. (45) Findings suggest that VV volumes may be an important biomarker for preterms with severe GMH-IVH.

With more preterm infants surviving, but with morbidity rates unchanging, the ultimate goal of this longitudinal study was to identify 3D cUS and fNIRS markers that predict intervention thresholds for infants with PHVD. Hence, we used 3D cUS and fNIRS to monitor preterm infants (≤ 32 weeks GA) with GMH-IVH. The overall hypothesis was that 3D cUS ventricle volume (VV) changes would be associated with regional changes in spontaneous functional connectivity (sFC). Improved understanding of the association between VV and sFC using bedside monitoring methods may improve the prediction of intervention thresholds in neonates with GMH-IVH. We had two main aims for this study. The first aim was to assess the association of ventricular morphology with sFC in preterm neonates assessed using 3D cUS and multichannel fNIRS. The second aim was to examine longitudinal assessments of 3D cUS ventricular morphology in neonates with severe GMH-IVH and PHVD who underwent CSF diversion procedures, and how they predict sFC.

## Methods

### Study Setting and patients

This prospective cohort study was conducted at the Level 3 Neonatal Intensive Care Unit (NICU) in Ontario, Canada, from January 1, 2021, to June 30, 2022. Families provided informed consent. Infants (≤ 32 weeks’ GA) receive a routine cUS within one week of age as part of standard care. (33) Study participants were infants ≤ 32 weeks’ GA, born at or referred to London Health Sciences Centre (LHSC) NICU, and admitted with a diagnosis of GMH-IVH, (46) made by the most responsible physician on the infant’s first routine 2D cUS. The Health Sciences Western University’s Research and Ethics Board approved the study.

### Data collection

#### Demographic and clinical data

Maternal and neonatal data were abstracted from electronic medical records and charts. For each infant, we collected the following data: *Antenatal and perinatal*: maternal age, medical conditions (hypertension, diabetes mellitus), place of birth (inborn vs outborn), corticosteroids administration (complete course vs incomplete course or none); mode of delivery; gestational age at birth; birth weight; sex; invasive ventilation at birth, delayed cord clamping, cord pH, Apgar score at 1 and 5 min. *Prematurity-related clinical morbidities*: patent ductus arteriosus (PDA) requiring treatment, sepsis (culture positive), hypotension requiring inotropes, bronchopulmonary dysplasia (the need for supplemental oxygen at 36 weeks’ postmenstrual age), retinopathy of prematurity (ROP), periventricular leukomalacia (PVL). *Ventricular measurements and fNIRS data*: 2D cUS GMH-IVH staging (right, left), 3D cUS ventricle volumes (left, right and total), and postnatal course during each measurement; resting state fNIRS data. *For infants with PHVD, before and after each tCSF (VT or EVD) procedure*: head circumference, weight, hemoglobin, ongoing respiratory support (invasive vs non-invasive ventilation or room air); tap volume; the need for VP shunt; brain MRI for those infants who went on to have one.

#### 3D cUS system and ventricle volume acquisition

We used a 3D cUS system to image the lateral ventricles attached to a clinical 2D cUS machine (HDI 5000, Philips, Bothel WA) with an appropriate conventional probe (C8-5, Philips, Bothel WA). (47, 48) The 3D cUS system comprises a handheld motorized device housing a 2D cUS probe (Figure 1). The probe was placed on the infant’s anterior fontanelle (the soft part of the brain) with the infant lying supine. After initiating the scan, the device tilted the probe on an axis at the probe tip. 2D cUS images were acquired into a computer via a digital frame grabber (Epiphan DVI2USB 3.0) and reconstructed into a 3D image. (44) Total bedside scan lasted 5-10 minutes, accounting for infant movement and large ventricles. We manually segmented cUS images using in-house developed software. Segmentation of the lateral ventricles was performed in two sagittal and coronal planes by trained researchers (LMK, ML, BK and SA) and verified by a Paediatric Neurosurgeon (SdR). The software created a 3D image of the lateral ventricles, from which a ventricle volume (VV) measurement was provided. (49) This system has been validated for geometric validity and volumetric measurements. (48) Depending on VV, the infants underwent sequential 3D cUS, weekly or biweekly, until discharge or term equivalent age (TEA), whichever came first.

**Figure 1:**
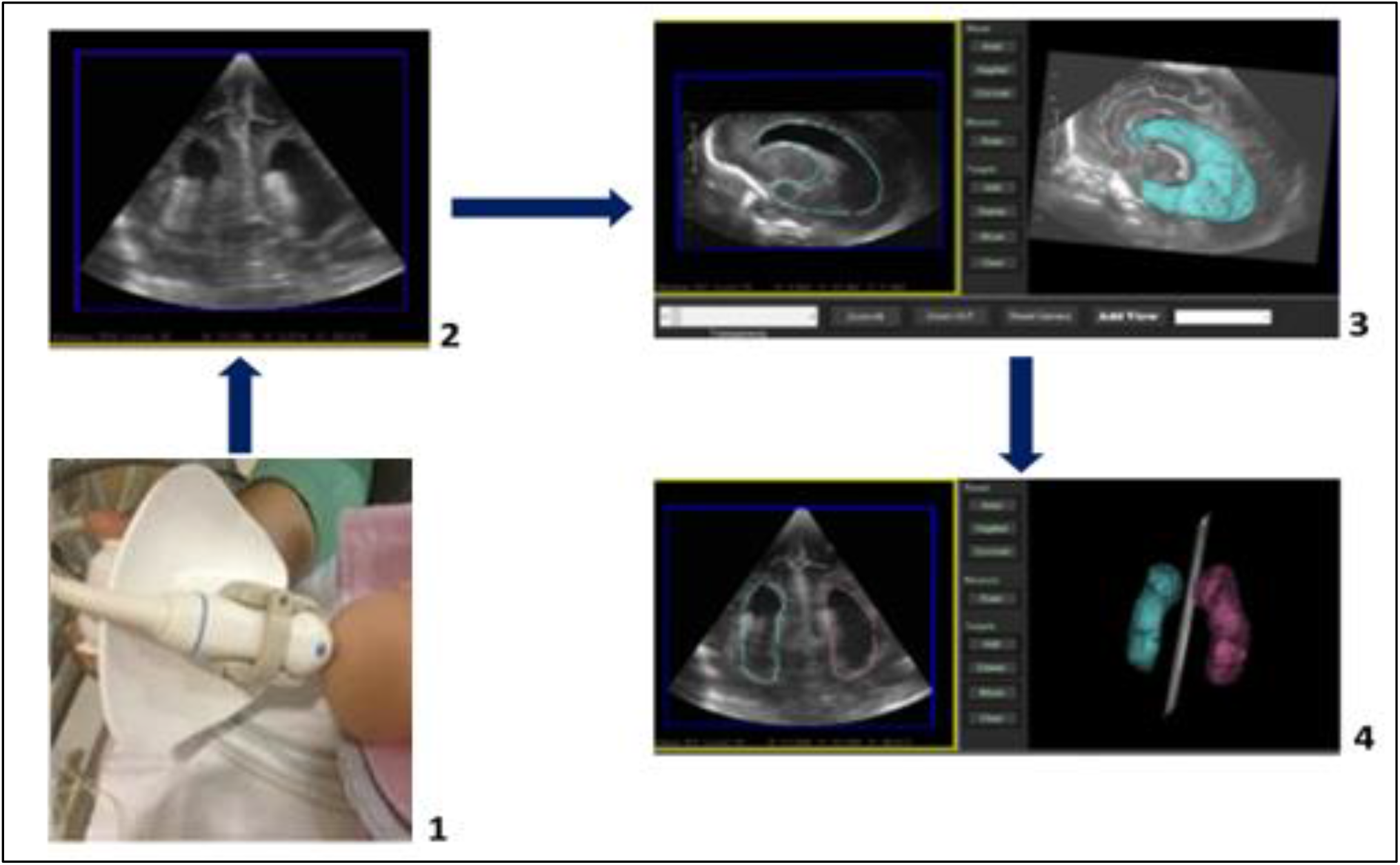
Three-dimensional cUS acquisition process: 1) Manikin head, with US probe placed on anterior fontanelle. 2) cUS image 3) lateral ventricle segmentation 4) segmented ventricles (left – pink; right – blue), from which a VV is provided.

#### fNIRS data acquisition

While infants lay in the incubator, we acquired fNIRS signals using a multichannel NIRSport2 system (NIRStar Software v14.0, NIRx Medical Technologies LLC, Berlin, Germany) at a sampling rate of 10Hz, (Figure 2). We inserted eight sources and eight detectors into predefined cap areas, with ten channels (with 22mm separation) defined for each hemisphere (20 total). The optodes were placed over the prefrontal and sensorimotor regions. Sixteen early recordings of the total 182 recordings used a slightly different montage so all datasets have been merged to contain 20/24 possible channels (Supplementary figure 1). Each light source contains two LEDs that emit at 760 nm and 850 nm. According to international standards, the fNIRS cap was positioned on the infant’s head. (50) Resting state fNIRS were acquired for a minimum of 2.5 minutes and mean of 6.8 minutes. (51)

**Figure 2:**
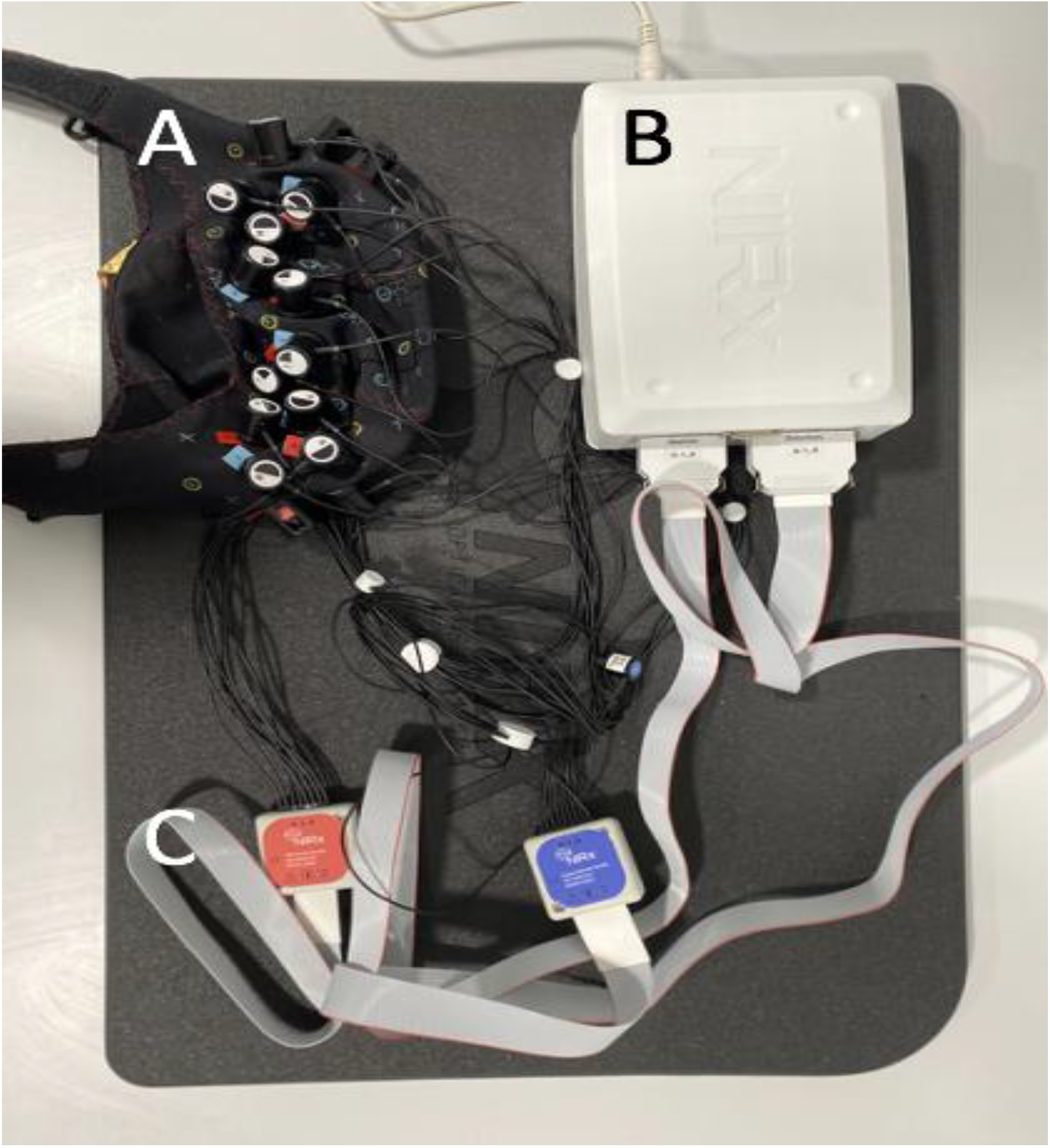
fNIRS acquisition system. A. fNIRS cap. B. NIRSport2 system. C, Optodes

#### Measurements’ sequence and timing

We performed the fNIRS recording first, followed by 3D cUS at the infant’s bedside. Infants with severe GMH-IVH who demonstrated worsening PHVD, requiring a VT or EVD, underwent 3D cUS and fNIRS before and after the intervention (on the same day). Hence, all subjects had multiple 3D cUS VV and fNIRS measurements.

#### fNIRS patient exclusions and subgroups

Three patients with fewer than two valid sessions were excluded. The remaining patients were divided into two groups: those who did not undergo CSF diversion procedures (no CSF diversion group) and those who did require CSF diversion procedures (CSF diversion group). The data from these groups were analyzed separately given the known changes in brain morphology with CSF diversion procedures.

#### fNIRS subsample selection

Each recording was reduced to the contiguous 2.5-minute subsample (51) with the highest data quality using in-house scripts and metrics adapted from QT-NIRS (also known as NIRSPlot). (73) First, abrupt motion spikes were interpolated using an established spline-based method adapted from Homer2. (52, 53) Scalp coupling index and peak spectral power were calculated in 5-second windows with 50% overlap for a cardiac range of 90 to 210 beats per minute. Selections were then made such that scalp coupling index and peak spectral power were maximized while segments with signal dropout (e.g., from optodes lifting off the scalp) were avoided.

#### fNIRS channel exclusions

We excluded channels within each dataset where cardiac pulsation was absent, or other issues were manually identified. Cardiac pulsation was detected using a novel method that combines the scalp coupling index with patterns in the frequency domain. Manual corrections were applied based on visual inspection. 14.8% of channels were excluded due to absent cardiac pulsation, excessive motion, and/or signal dropout. An additional 7.3% of channels were excluded due to a hardware malfunction in source 8 that impacted 3 channels in the right hemisphere. These exclusions resulted in variable channel validity across sessions and between patients. Channels that did not meet a minimum sample size threshold (10/23 or 5/7) were indicated as “low confidence” and were excluded from analyses.

#### fNIRS data preprocessing

A combination of Brain AnalyzIR Toolbox (74), Homer2 (52), and in-house MATLAB scripts were used for fNIRS data processing. The data were converted to optical density and then to HbO and HbR using the Brain AnalyzIR Toolbox. (74) Age-appropriate partial pathlength factors (0.1063 for 760nm and 0.0845 for 850nm) were used in the modified Beer-Lambert Law. (54) Additionally, channel lengths were scaled to the cap size used during each session. The data were then converted to HbT (HbO+HbR) which has been shown to have improved reproducibility across sessions and subjects. (53) Nuisance regression was performed which applied temporal filtering (roughly equivalent to a bandpass around 0.004 Hz to 0.09 Hz) and was also the primary means of motion correction. (55) The final data were resampled to 1 Hz before calculating correlations (see supplementary figure 2 and 3).

#### Spontaneous functional connectivity

The Brain AnalyzIR Toolbox was used to calculate sFC using a robust correlation method. (74) Prewhitening was not applied. (56) To validate the sFC, group summary figures were generated as follows: correlations were first averaged across sessions to yield one mean value per patient channel pair; a single-sample t-test was performed on each channel pair to evaluate sFC ≠ 0; the resulting t-map was drawn over the montage.

#### Relating spontaneous functional connectivity to ventricle volumes

We used absolute sFC correlations (|sFC|) because we expected impairments to involve reductions in the positive correlations and increases in the anti-correlations (i.e., all correlations trending towards zero, a decrease in magnitude). The relation of |sFC| to VV was assumed to be linear and evaluated by estimating the slope of |sFC| / VV. We did not use correlation because several patients had as few as two sessions. The |sFC| / VV slope was estimated using three incremental methods. All three methods were calculated independently for each channel pair using VV from the corresponding hemisphere/ventricle. The first method used simple linear regression to estimate each patient’s slope. The second method used linear mixed-effects modelling (LME), which was less sensitive to outlier patients. The third method used LME and additionally accounted for changes in sFC and VV across GA.

A t-map was generated for each method for |sFC| / VV ≠ 0. Positive values indicate where |sFC| increases/decreases with VV, and negative values indicate where the relation is inverted (e.g., decreasing |sFC| as VV increases). For a broad comparison, we performed a subsequent t-test (|sFC| / VV ≠ 0) on the group-level slopes from each channel-pair (separate for each hemisphere, for each method). This test indicates where significant trends occurred at the hemisphere level.

#### Longitudinal case studies in patients requiring CSF diversion procedures

For each case study, we present |sFC| and VV sessions across GA and indicated the times of each diversion procedure. As a middle ground between presenting every channel pair and collapsing to hemispheres, |sFC| are presented from four fully symmetrical functional clusters identified in the sFC validation t-maps. Each cluster contains four channel pairs derived from five channels. (Supplementary Figure 5).

### Statistical analysis

SPSS v.28 (IBM Corp., Armonk, NY, USA) was used for all statistical analyses. To address our first aim, we examined the association between VV and sFC, by estimating the linear relation (slope) of |sFC| across VV. To address our second aim, we additionally investigated |sFC| and VV across time as individual case studies only in infants who underwent CSF diversion procedures. Models were adjusted for relevant covariates and confounding variables. As we had a single hypothesis and did not make any direct comparisons across the groups or methods, an alpha level s of 0.05 was considered statistically significant.

## Results

### Study population

During the study period, 43 preterm neonates (GA ≤ 32 weeks, with GMH-IVH) were eligible for 3D cUS and fNIRS monitoring. We excluded two neonates (extremely low for gestation (ELGA), < 26 weeks) following their death during the first week of life. The neonates died after a decision for palliative care in the NICU and before data acquisition. The decision to redirect care was based on the degree of prematurity, the severity of GMH-IVH and other prematurity-related complications. We excluded seven neonates because they lacked fNIRS measurements and in another neonate, it was not possible to obtain 3D cUS measurements. In addition, three patients with fewer than two viable sessions were excluded. Thus, our study population consisted of 30 preterm infants (mean GA 26.6 weeks, SD 2.66; mean birth weight 1,002.4g, SD 362.681) with 3D cUS and fNIRS measurements. Figure 3 is the study flow diagram. The remaining 30 patients were divided into two groups: those who did not undergo CSF diversion procedures (n=23, “No CSF Diversion” group) and those who required CSF diversion procedures (n=7, “CSF Diversion” group).

**Figure 3:**
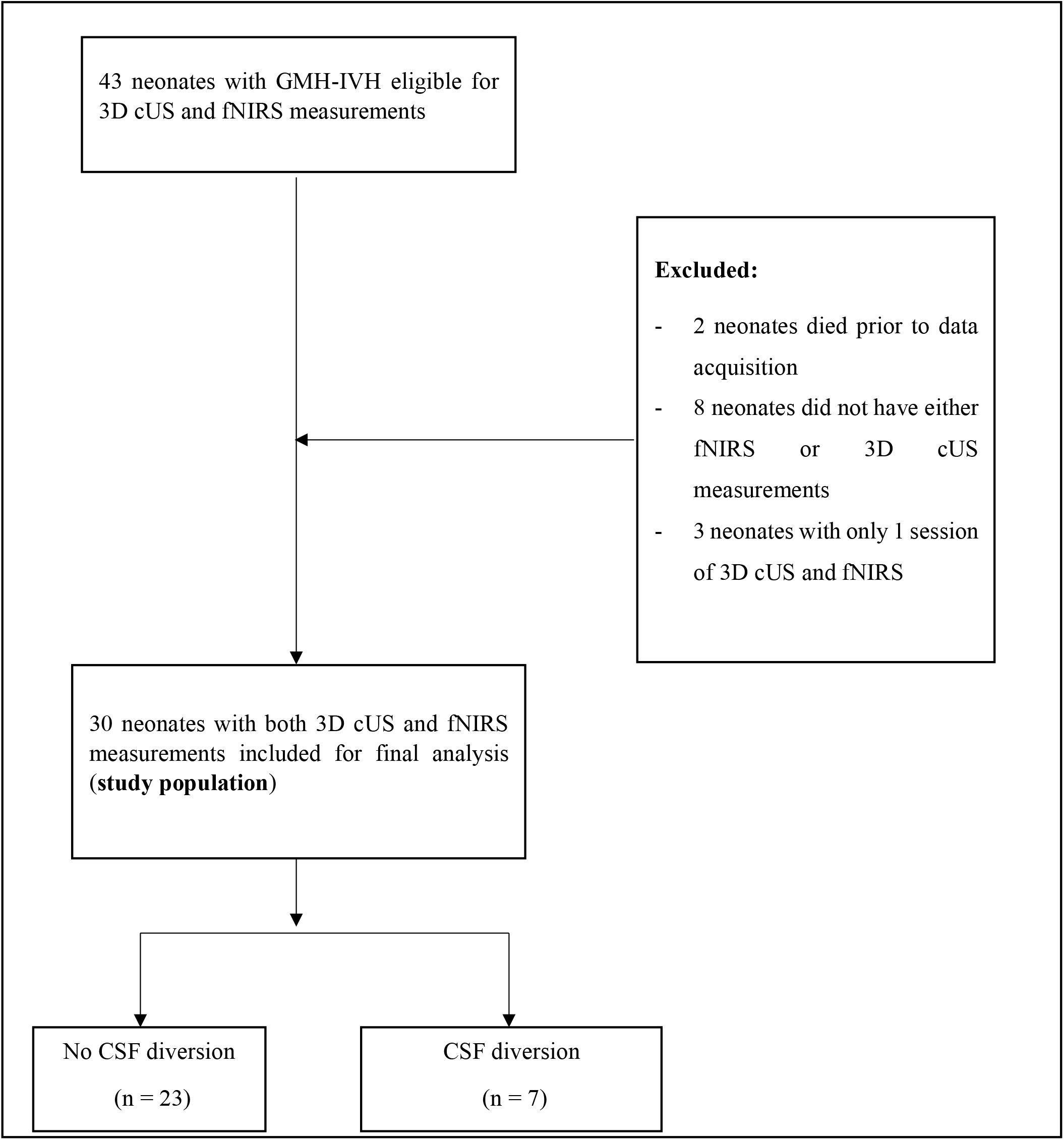
Flow diagram

Most infants were born at LHSC (24, 80.0%), and 16 (53.3%) were female. Only twelve mothers (40.0%) received a complete course of antenatal corticosteroids. The majority of mothers presented in spontaneous preterm labour and hence did not benefit from antenatal corticosteroid interventions. A higher proportion of infants (20, 66.7%) remained intubated during the first week of life. Out of the 30 neonates enrolled, 19 (63.3%) had mild GMH-IVH, while 11 (36.7%) had severe GMH-IVH. Severe GMH-IVH was defined as GMH-IVH grade III or PHVI. Seven patients (24.2%) with severe GMH-IVH had temporizing CSF procedures (ranging from 2 to >15), while three infants (10.0%) had a VP shunt inserted before discharge. In addition, the majority of the study participants had prematurity-related complications: hemodynamically significant patent ductus arteriosus (17, 56.7%), clinical or culture-proven sepsis (25, 83.3%), severe retinopathy of prematurity (18, 60.0%). Table 1 shows demographic characteristics and clinical data. The “CSF Diversion” group had a mean GA of 26.88 weeks and a mean birth weight of 1143.25g. Five out of seven neonates (62.5%) were outborn. Of the seven neonates, 2 (28.6%) had grade II IVH, 2 (28.6%) had grade III IVH, and 3 (42.9%) had PHVI.

**Table 1:** Demographic and clinical data of the study cohort (n = 30)

| Characteristics | (n = 30) |
| --- | --- |
| **Neonatal characteristics** |  |
| Gestational age at birth (wk) | 26.6 ± 0.95 (23-32) SD. 2,66 |
| Birth weight (g) | 1,002.37 ± 129.78, (590 - 1910), SD = 362.681 |
| Multiple births (>2) | 12 (40.0) |
| Male sex | 14 (46.7) |
| Apgar 5 min | 6.3 ± 0.84 (1-9) |
| Inborn | 24 (80.0) |
| Cesarean section delivery | 11 (36.7) |
| **Maternal characteristics** |  |
| Antenatal corticosteroids (complete course) | 12 (40.0) |
| Maternal age | 31.03 ± 2.31 (17-46) |
| **Neonatal complications** |  |
| Invasive ventilation > 1 week | 20 (66.7) |
| hsPDA | 17 (57.6) |
| **GMH- IVH grade** |  |
|  | **Grade I:** 7 (23.3) |
|  | **Grade II:** 12 (40.0) |
|  | **Grade III:** 6 (20.0) |
|  | **Grade IV:** 5 (16.7) |
| Culture-positive or clinical sepsis | 25 (83.3) |
| Hypotension, inotrope use | 10 (33.3) |
| BPD | 12 (40.0) |
| Severe ROP | 18 (60.0) |
| PVL | 12 (40.0) |
| tCSF diversion procedure | 7 (23.3) |
| VP shunt | 3 (10.0) |
| Death before discharge | 1 (3.3) |

**RDS**, respiratory distress syndrome; **hsPDA** - hemodynamically significant patent ductus arteriosus, **ROP** - retinopathy of prematurity; **PVL** - periventricular leukomalacia; **tCSF** - temporizing cerebrospinal fluid diversion procedure; **VP** shunt - ventriculoperitoneal shunt

### Validation of spontaneous functional connectivity

It was important to first establish the presence of sFC and identify any existing trends. Figure 4 represents t-maps for both group-level sFC (top row represents the group that did not require CSF diversion, mean 3.6522 SD, 1.8243, and the bottom row – CSF diversion group, mean 9.5714, SD 4.3150). Both t-maps include clusters of adjacent channels that are positively correlated, anti-correlations between those clusters, and many significant anti-correlations between hemispheres (see supplementary figure 4 for anti-correlations between hemispheres). These are expected patterns and do not raise any concerns moving forward. Supplementary figures 6 summarizes number of sessions per patient in both groups.

**Figure 4:**
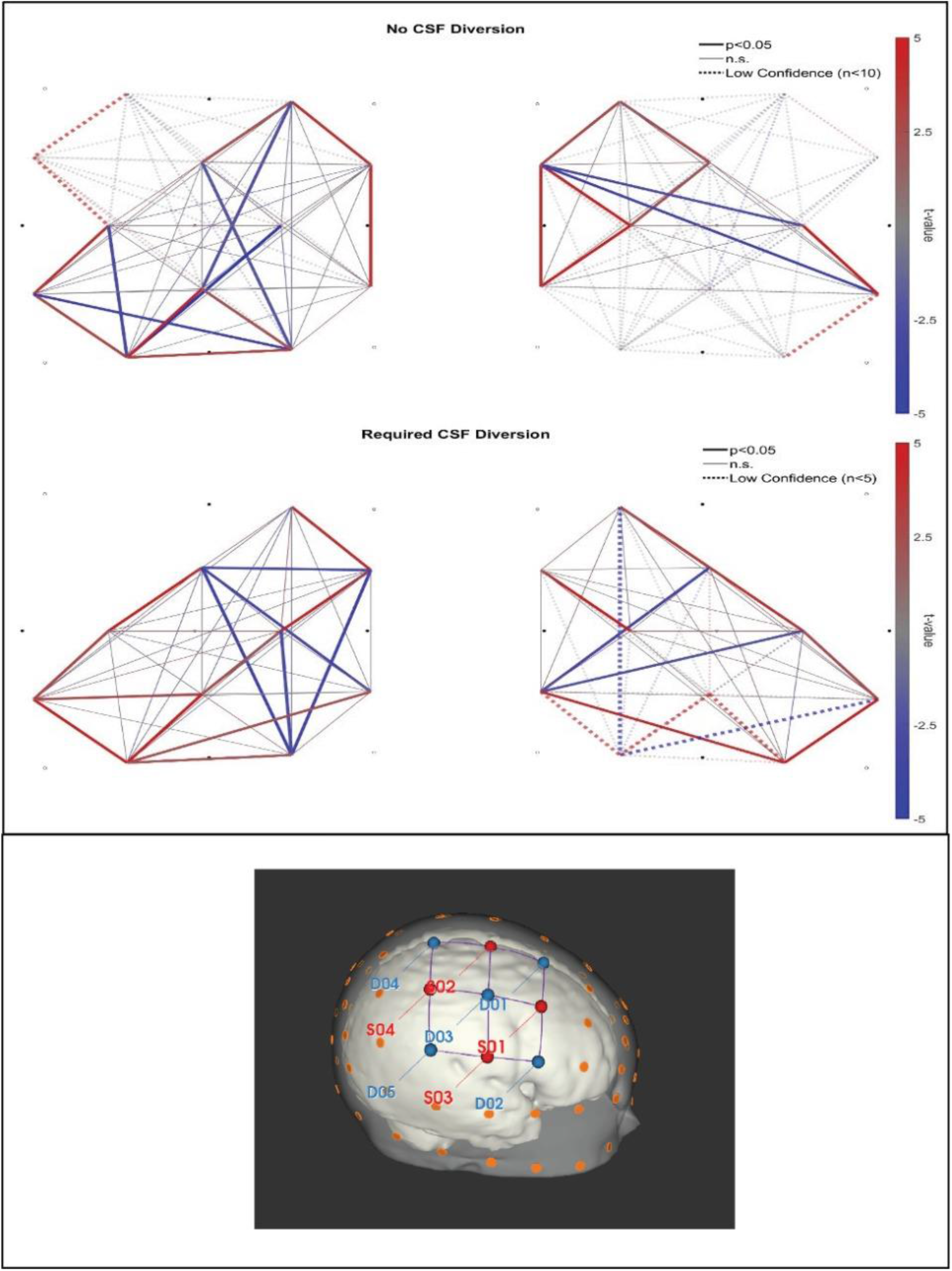
Group level sFC t-maps (top row, “No CSF Diversion group”, n = 23; bottom row, “Required CSF Diversion” group, n= 7). The left hemisphere is on the left and the right hemisphere is on the right. Included at the bottom is a montage of the neonatal head.

### Ventricular morphology and spontaneous Functional Connectivity

To assess our first aim, the association of ventricular morphology with sFC in preterm neonates assessed using 3D cUS and multichannel fNIRS was examined. In the “No CSF Diversion” group, the linear regression method produced a widespread negative |sFC| / VV (see Figure 5) that was significantly less than zero in the left hemisphere (-0.0729, p<0.001) and trending similarly in the right (-0.1030, n.s.). All results are outlined in Figure 6.

**Figure 5:**
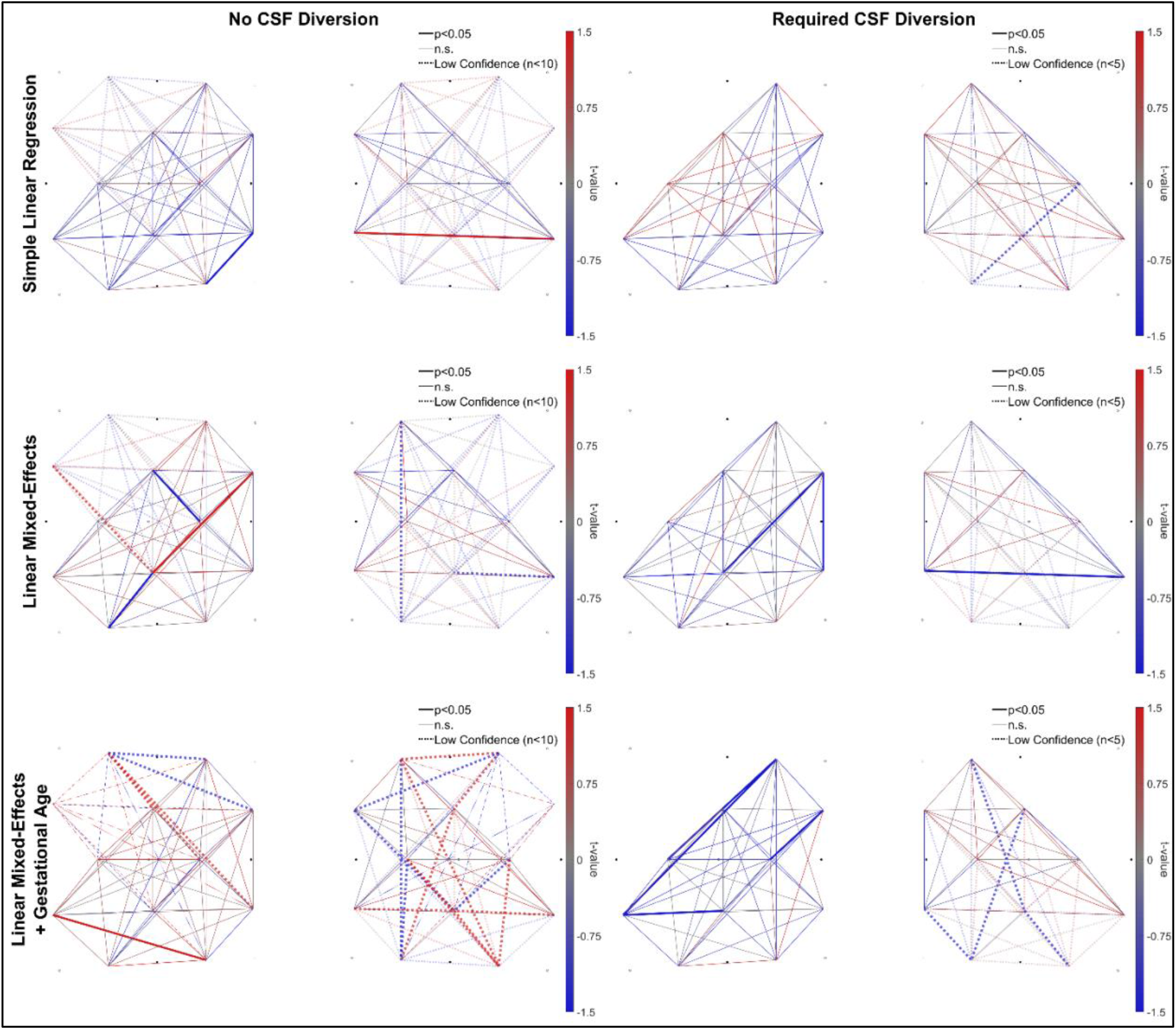
Group level |sFC|/VV t-maps from each slope method (left = “No CSF diversion” group, n = 23; right = “Requiring CSF diversion” group, n = 7); Rows: top = simple linear regression, middle = LME, bottom = LME + GA

**Figure 6:**
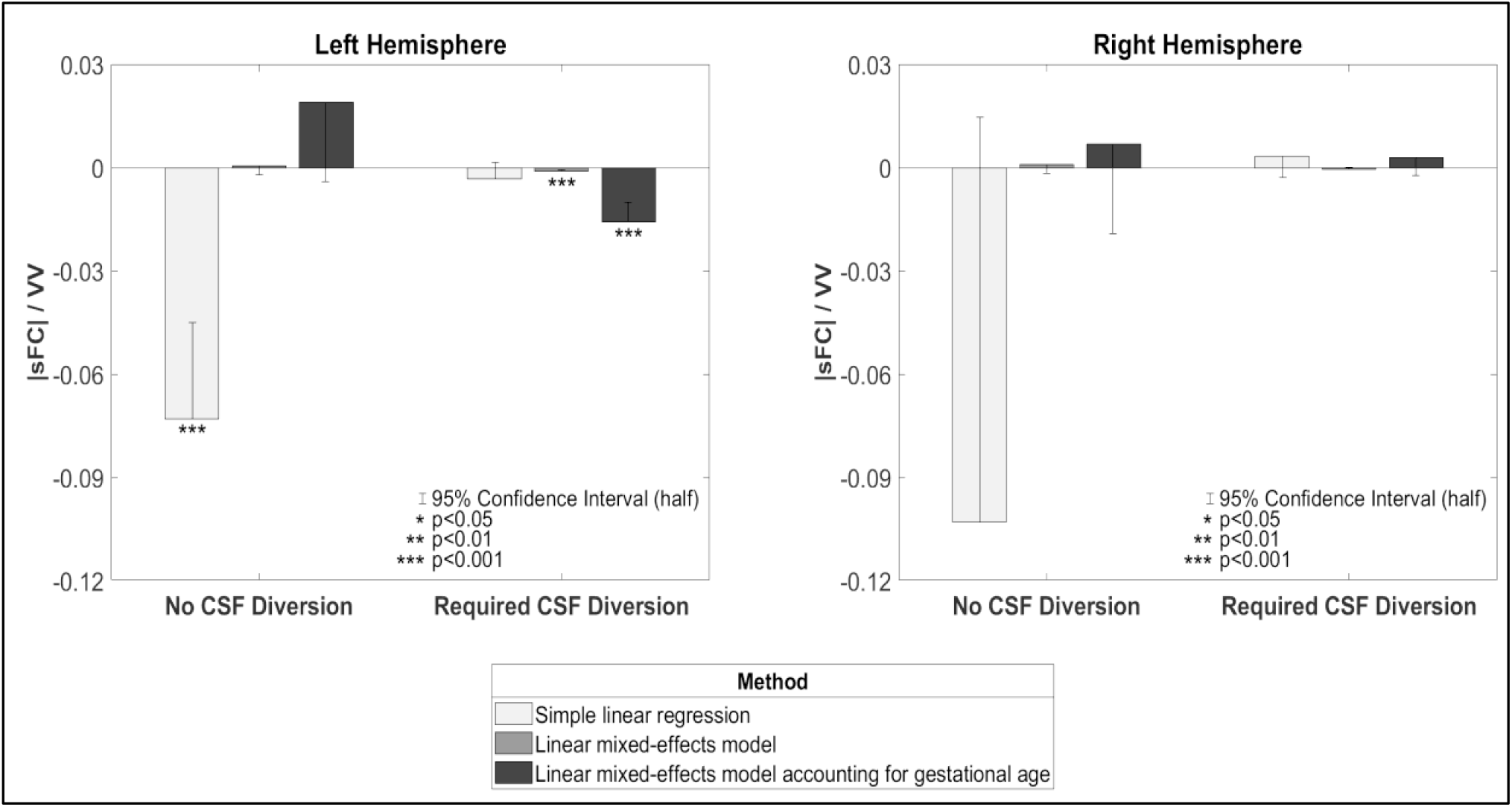
Hemispheric trends in group level |sFC|/VV from each slope method.

In the “Required CSF Diversion” group, |sFC| / VV trended negatively in the left hemisphere across linear regression (-0.0032, n.s.), LME (-0.0012, p<0.001), and LME additionally accounting for GA (-0.0159, p<0.001). Several mostly-adjacent channel pairs were independently significant in both LME results. However, the right hemisphere was inconsistent across all three methods.

To provide context to the results of |sFC| / VV, we performed a post-hoc investigation of possible trends in VV and |sFC| across GA in each group. The only significant finding was a decrease in VV across GA in the left hemisphere of the “No CSF Diversion” group (-0.3869 VV / Week, p<0.05), but this was not maintained when accounting for patient variability. Overall, there were no reliable main effects of GA on VV or |sFC| in this clinical population.

### Longitudinal assessments of 3D cUS ventricular morphology and sFC prediction

To address our second aim, concerning longitudinal assessments of 3D cUS ventricular morphology in neonates with GMH-IVH and PHVD, we examined how they covary with sFC. Two of seven case studies (not known to anyone outside research group) from participants requiring CSF diversion procedures are presented in Figure 7 (vertical gray lines indicate timing of CSF diversion). An inverse relation between |sFC| and VV is evident even when CSF is diverted. Furthermore, increases in |sFC| following CSF diversions were often observed shortly after each procedure. For all neonates who underwent CSF diversion, left, right and total VV decreased (VV decrease left, p<0.001, mean decrease -9.74, SD 7.76; VV decrease right p<0.001, mean decrease -10.20, SD 8.98; VV decrease total p<0.001, mean decrease -19.93, SD 12.23). All clusters/regions, except the left posterior, had a mean increase in |sFC| but not significant at the group-level (left anterior: |sFC| change p = 0.22, mean change = 0.04, SD 0.20; left posterior: |sFC| change, p = 0.51, mean change = -0.00, SD 0.27, right anterior: |sFC| change p = 0.10, mean change = 0.07, SD = 0.19; right posterior: |sFC| change p = 0.21, mean change = 0.06, SD 0.27).

**Figure 7:**
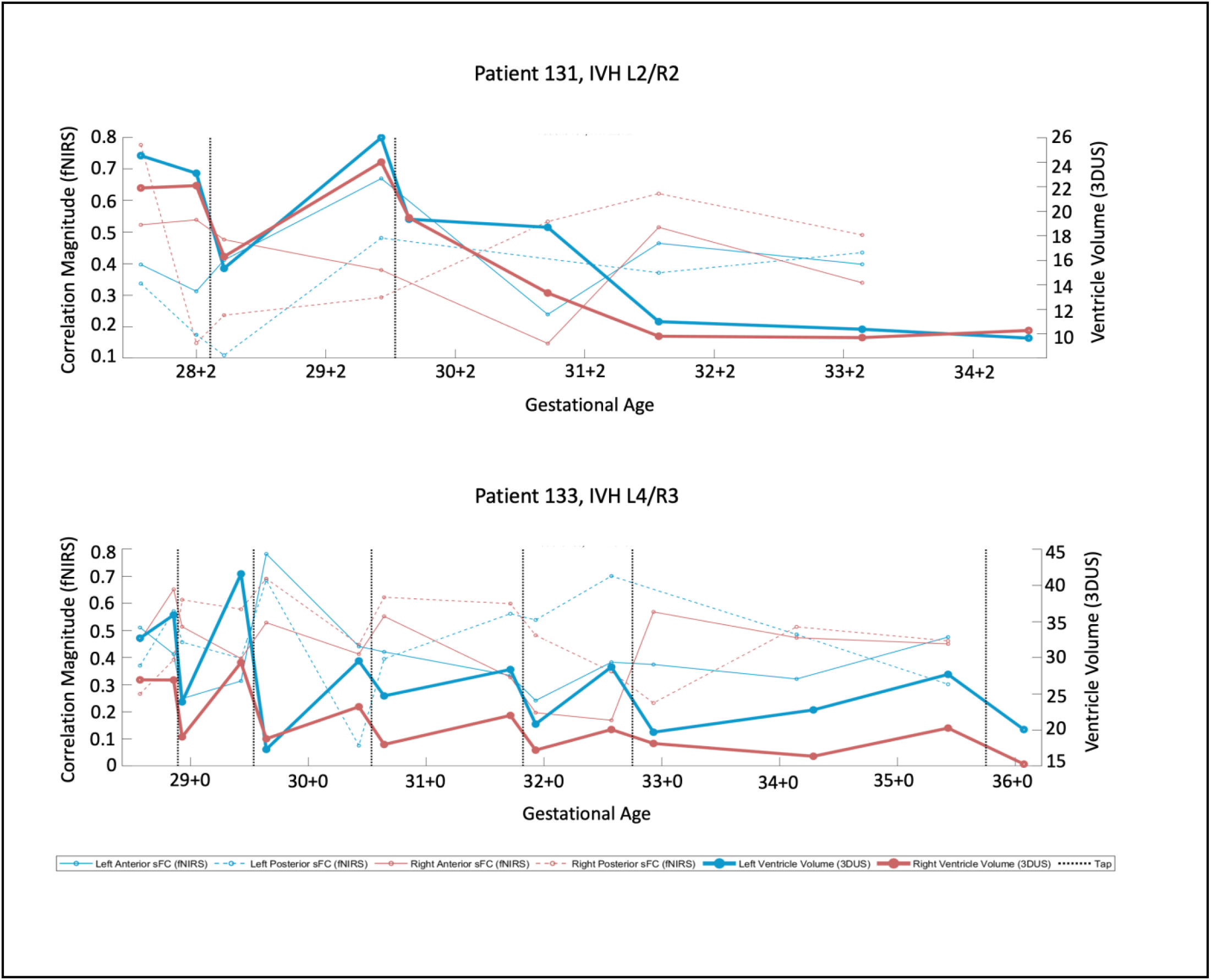
Two case studies from patients requiring CSF diversion. *Patient IDs 131 and 133 not known to anyone outside research group. *Vertical gray lines indicate CSF diversion*.

## Discussion

Despite obstetric and neonatal health improvements, GM-IVH remains unacceptably high in preterm neonates. This study used a combination of bedside tools - 3D cUS and fNIRS to monitor preterm infants with GMH-IVH from diagnosis until discharge. To our knowledge, no other study has incorporated these tools simultaneously. Our study aimed to determine if ventricular size in preterm infants with GMH-IVH was associated with changes in sFC. We observed an inverse relation between |sFC| and large VV: increases in VV were associated with decreases in |sFC|. Our findings of increased ventricular volume in preterm neonates and reduced fNIRS-based functional connectivity suggest that regional disruptions of ventricular size may impact the development of the underlying grey matter. (57) These results bear clinical significance given that the preterm neonate’s brain is rapidly growing and at risk for injury. (58, 59) Furthermore, severe GMH-IVH and PHVD have been linked to adverse ND outcomes. (40, 59)

In our first aim, in participants who did not have CSF diversion, the linear regression method produced a significant negative |sFC| / VV slope in the left hemisphere with a similar trend in the right, which indicates impaired sFC in preterm neonates with GMH-IVH. We also observed similar but more compelling results in the participants who required CSF diversion group: consistently negative across all the three methods used. The slopes in the “No CSF Diversion” group switched to positive under the LME-based methods, which suggests that most infants in this group had a positive relation (likely because most of these infants had mild GMH-IVH) and that the initial linear regression result was likely driven by the few negative outliers. Our findings suggest that increasing ventricular dilatation is associated with reduced |sFC| in preterm infants with GMH-IVH. To put this into context, the larger the ventricle, the more likely sFC is impaired. Our study findings are not surprising, given that the pathological and functional consequences of GMH-IVH depend on its severity. (57) Previous resting-state FC fMRI studies have reported similar findings, showing resting state networks development was affected depending on WMI severity and location. (60, 61) In addition, in a recent study, Tortora et al showed regional impairment of cortical and regional grey matter perfusion in preterm infants with mild GMH-IVH. (62)

As expected in our second aim, in the participants who required CSF diversion group, we observed elevated VV and reduced |sFC| right before the CSF diversion procedures. Conversely, after the diversion, VV decreased, and |sFC| often increased, although the results were not statistically significant at the group level likely due to a small sample size. Observed increases in |sFC| were observed shortly after each CSF diversion procedure, possibly correlating to a decrease in ICP. Our findings are consistent with others. In their study of 9 preterm infants, Norooz et al. reported an increase in the mean regional cerebral oxygen saturation (rcSO2) value after decompression (42.6 ± 12.9% before vs 55 ± 12.2% after decompression). (63) Our findings are also comparable to Kochan et al. Their study, which included 20 VLBW premature infants with GMH-IVH, demonstrated that ventricular dilatation was associated with lower cSO2, suggesting a decrease in cerebral perfusion. (64) Other studies in preterm infants with PHVD requiring CSF diversion procedures have consistently reported similar findings. (65-67) Similarly, Rajaram et al. and McLachlan et al. showed that cerebral blood flow increased after ventricular taps, further reinforcing our sFC trends. (67, 68)

Our findings show that fNIRS may provide additional clinical information, particularly in preterm infants with PHVD, to help determine the optimal time for CSF diversion. While the definitive treatment of PHVD is VP shunt placement to divert CSF, this procedure is often delayed. During this delay, temporizing procedures are done to decrease ICP. However, a prolonged increase in ICP, can lead to brain damage that can impact both periventricular WM and cerebral grey matter. Furthermore, the timings of the temporizing CSF diversion procedures mentioned above are unpredictable and rely on subjective clinical signs, symptoms and 2D cUS measurements. fNIRS measurements, therefore, have the potential to aid in timely decision-making.

Our study has some strengths that are worth highlighting. A key strength of this study was the serial measurements. To date, 2D cUS remains the neuroimaging modality of choice for bedside GMH-IVH diagnosis. Current guidelines recommend the first 2D cUS by the first week of life to screen for GMH-IVH. After that, repeat cUS is completed at 4-6 weeks of life to screen for complications, mainly periventricular leukomalacia. Recent evidence recommends more frequent screening, particularly amongst very preterm infants, to help identify these complications and for timely intervention for those needing it. (69) Moreover, timely diagnosis of these complications provides a window of opportunity to have not only meaningful conversations with parents regarding prognosis but also an early opportunity for intervention for the affected infants. (70) Another strength of our study was the demonstration of functional connectivity in preterm neonates using multichannel fNIRS, enabling us to cover multiple brain areas simultaneously. Lastly, our NICU and Paediatric Neurosurgery team have incorporated 3D cUS for monitoring preterm infants with GMH-IVH (44, 45, 47, 48) for the past decade. Our current 3D cUS is semi-automated, requiring time to segment the lateral ventricles. For 3D cUS to play a role in diagnosing and monitoring PHVD in other clinical settings, an efficient and fully automated segmentation algorithm is crucial. Several other groups have adapted this technique, given its reliability in VV estimation. (71, 72) Nevertheless, we still achieved reliable VV estimates using our current system.

Some limitations of this study need to be acknowledged. First, the variability in the number of study measurements each participant had led to variability in the results. Our analysis showed a more robust and reliable sFC pattern when the study participants had three or more sessions. More sessions for each patient and fewer missing/excluded data would have enabled us to use more robust connectivity methods such as graph theory. Despite this, we still observed meaningful sFC patterns. Second, given the limited period, including follow-up data from this cohort was not possible. Morbidity is substantial in preterm infants with severe GMH-IVH and PHVD. Grade III and PHVI have been linked to CP, neurosensory problems and cognitive impairments; hence, it is only fitting that these outcomes are reported. Participants from this study are currently undergoing follow-up in the developmental follow-up and Paediatric Neurosurgery clinics. The neonates’ developmental outcomes will provide additional information regarding the clinical application of these tools and their implications. Third, the right hemisphere was inconsistent across linear regression in the “No CSF Diversion” group. This inconsistent finding could be attributed to a poor signal in one of the sources during the initial data acquisition period that may have influenced the results in the right hemisphere. Another plausible reason could be heterogeneity in our study participants (63.6% had mild GMH-IVH). Future prospective studies should examine larger populations of infants with all grades of GMH-IVH.

We included a model with gestational age because it is reasonable to expect an increase in both VV and |sFC| across GA and we wanted to investigate their relation that is separate from this trend. However, we did not observe any reliable main effects of GA, even in the milder “No CSF Diversion” group. It is likely that these effects were present but could not be detected due to a combination of low sample size and the noise introduced by IVH-related fluctuations in VV and |sFC|. In addition, these findings are not surprising given that with mild GMH-IVH, we expect to see the resolution of the bleeds with time and hence restoration of sFC. Previous studies have shown the maturation of sFC with advancing GA. Further studies are required to determine if GA should be included in the model.

Other things to consider are poor spatial resolution and limited penetration depth (especially in preterm infants with dependent scalp oedema). Such issues, however, are not uncommon in fNIRS studies, particularly in the very preterm population where the fit of the caps may be insufficient to make proper contact with the scalp. Therefore, future studies with larger cohorts are needed to confirm our results.

## Conclusion

In conclusion, our study shows that 3D cUS and fNIRS are promising bedside tools for monitoring GMH-IVH and PHVD in preterm infants, providing complementary information about the infant’s structure and function. Future research should explore the potential role of lateral ventricle volumes and spontaneous functional connectivity in the diagnostic and therapeutic approach to PHVD.

## Data Availability

The data that support the findings of this study are available from the corresponding author upon reasonable request.

## Acknowledgements

We thank NICU families whose consent and data made this study possible. We are grateful to all NICU staff and Paediatric Neurosurgery Residents for their immense help with this study.

## Authors contributions

LMK, KS, SdR and EGD conceptualized the study, designed and drafted the manuscript; LMK, ML, BK, and SA acquired data; KS and LMK analyzed data, and LMK, KS, KSL, SdR, EGD critically revised the article for important intellectual content.

