## Supplementary figures 1-6 for "Three-dimensional cranial ultrasound and functional near infrared spectroscopy for bedside monitoring of intraventricular hemorrhage in preterm neonates"

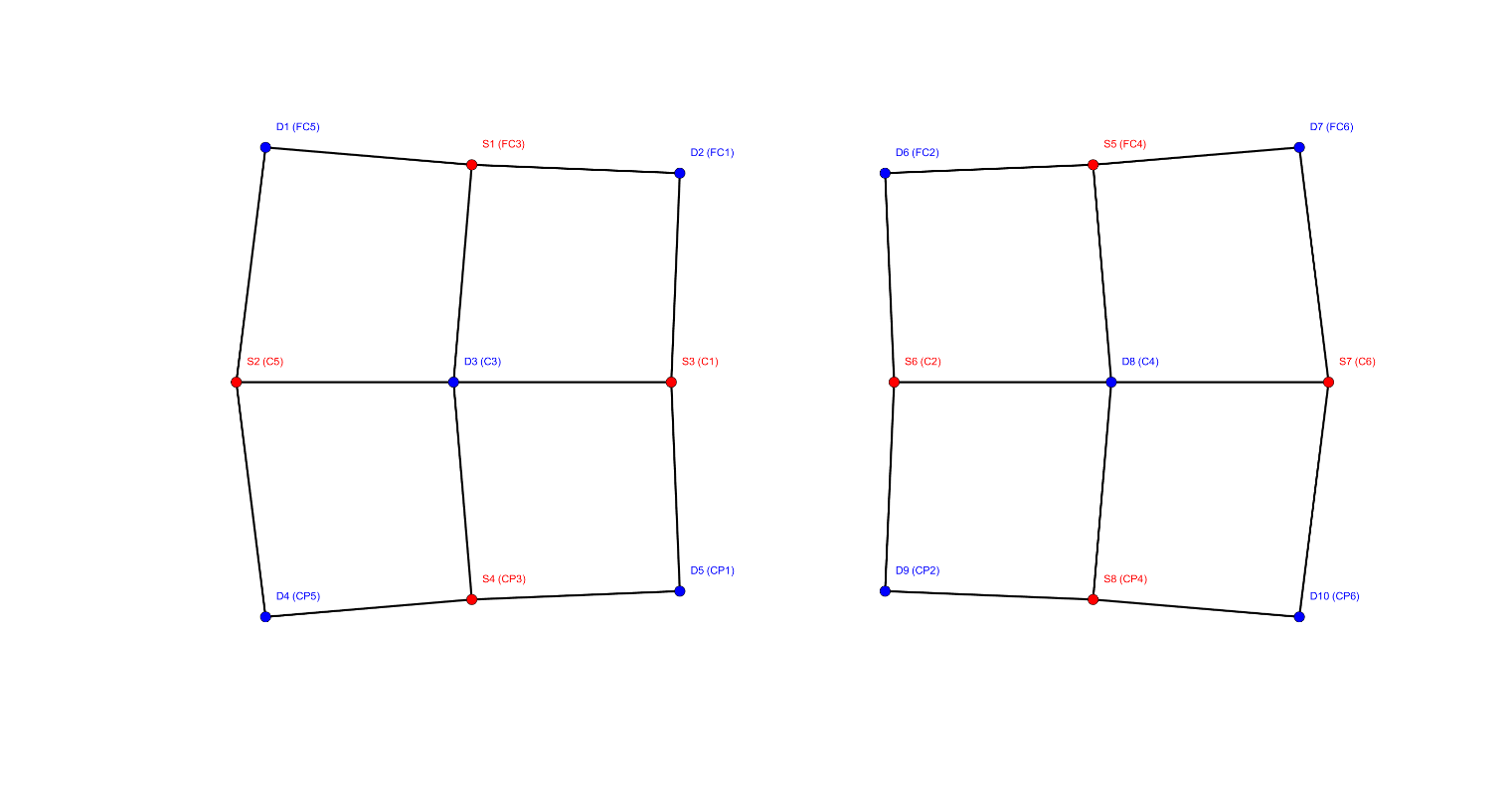

***Supplementary Figure 1:*** *Montage*

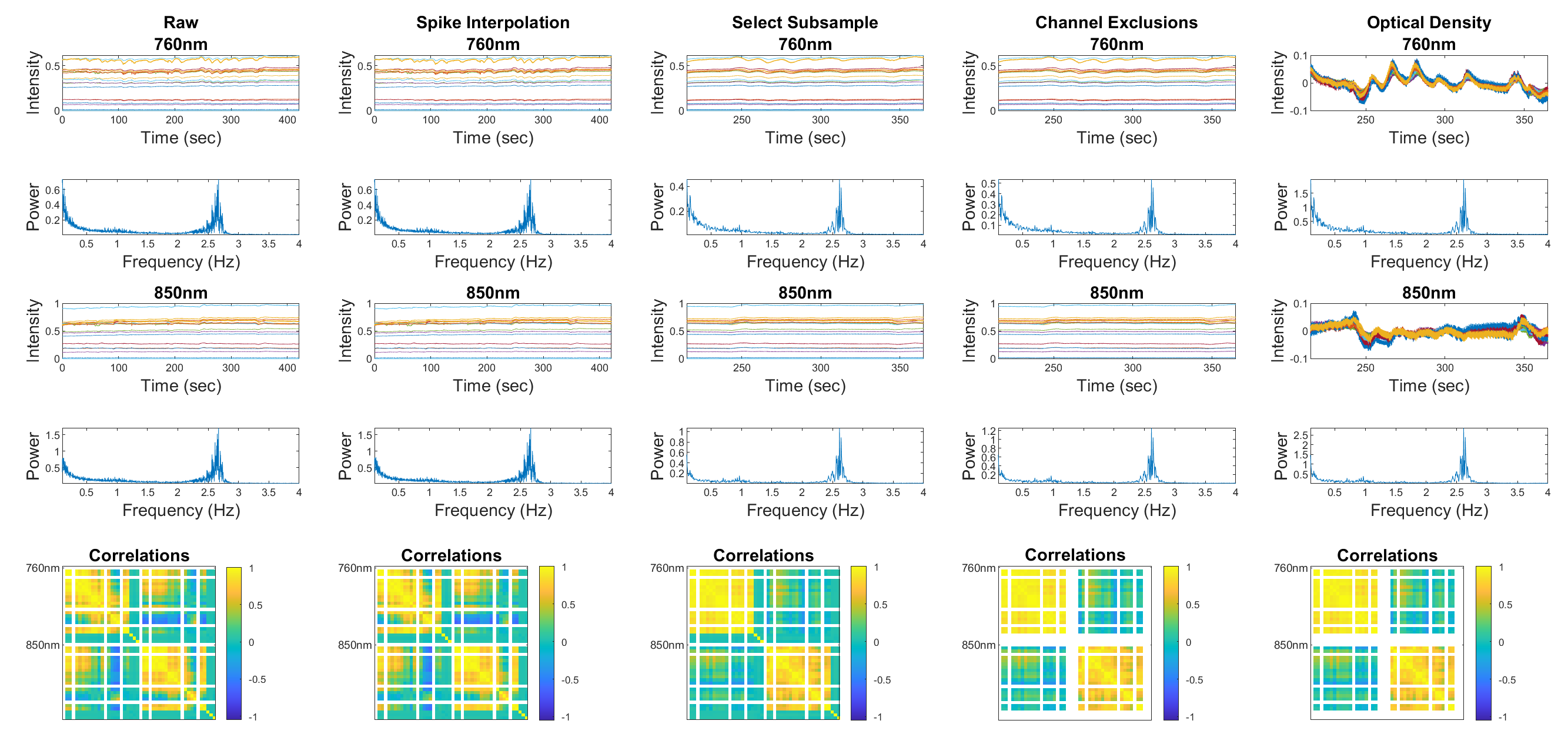

***Supplementary Figure 2:*** *Preprocessing pipeline, from acquisition of raw data to optical density*

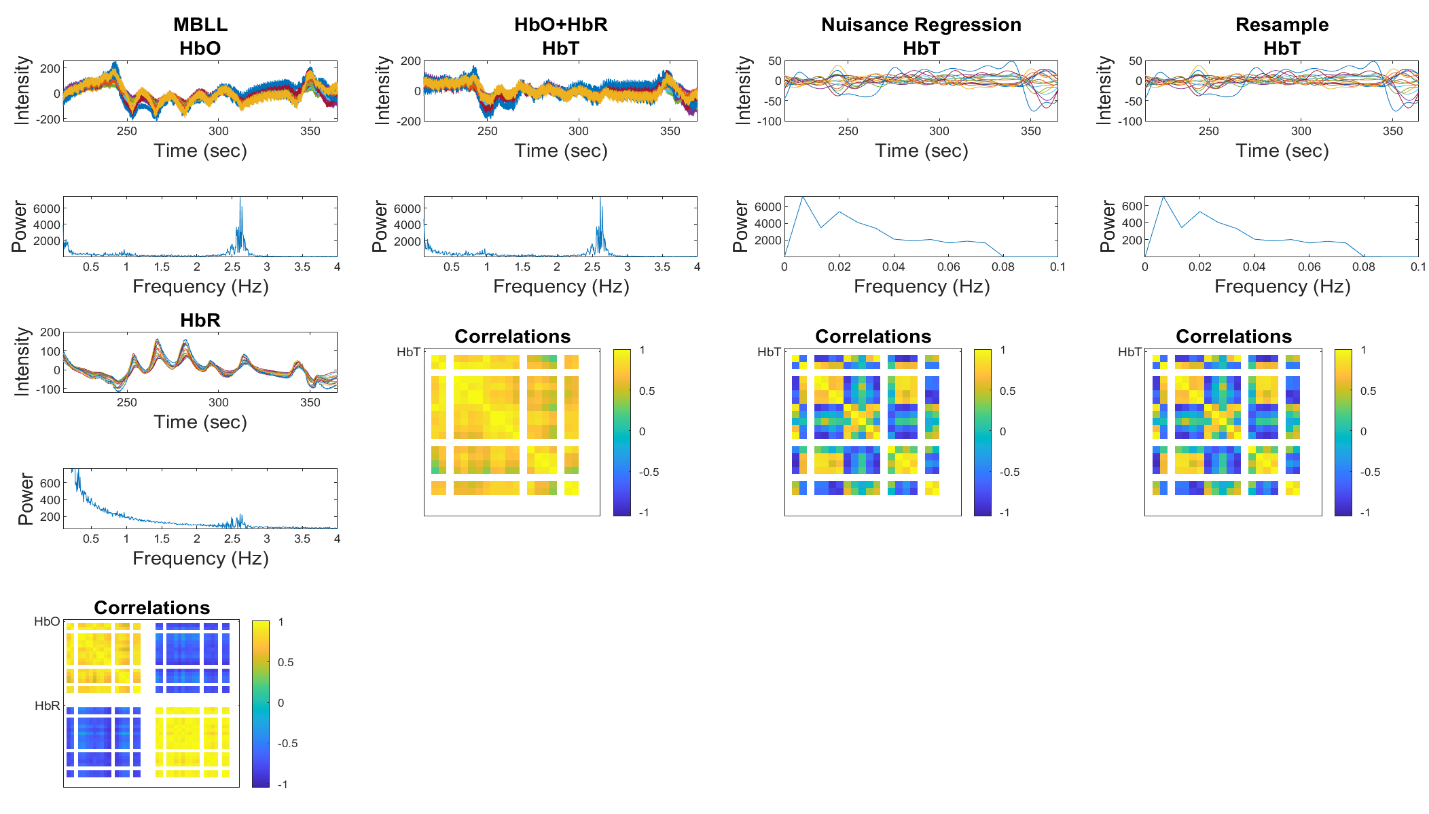

***Supplementary Figure 3****: Preprocessing pipeline (continued)*

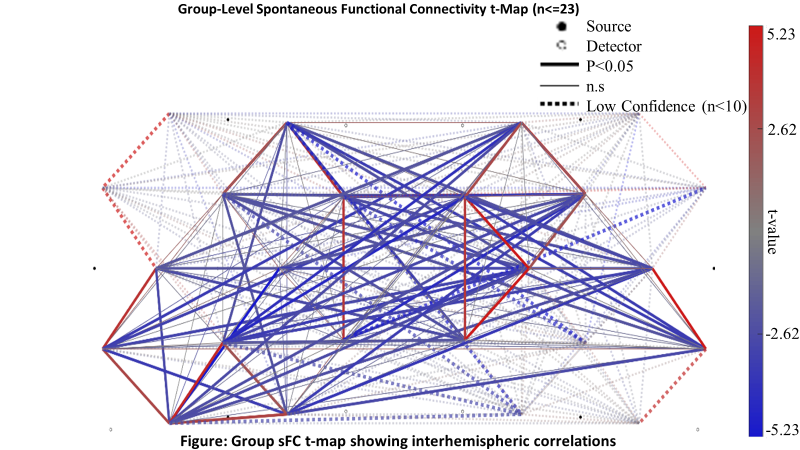

***Supplementary Figure 4:*** *Group t-map showing interhemispheric correlations*

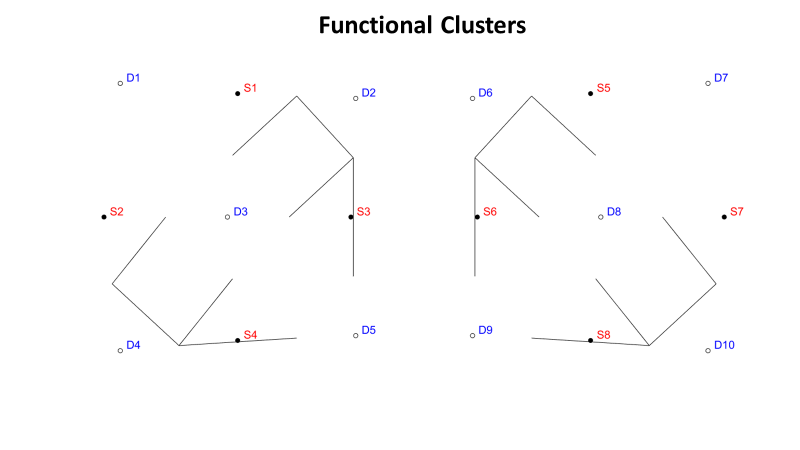

***Supplementary Figure 5****: Four functional clusters*

Number of sessions per subject in "No CSF Diversion Group" (n = 23)

| Number of sessions | Subjects |
| --- | --- |
| 2 | 9 |
| 3-5 | 9 |
| > 5 | 5 |

Number of Sessions per Subjects in "CSF Diversion Group" (n = 7)

| Number of Sessions | Subjects |
| --- | --- |
| < 5 | 2 |
| 5-10 | 1 |
| > 10 | 4 |

**Supplementary Figure 6:** Figure showing number of sessions per patient in both groups
